# Selection bias in reporting of median waiting times in organ transplantation

**DOI:** 10.1101/2023.12.13.23299859

**Authors:** Simon Schwab, Andreas Elmer, Daniel Sidler, Lisa Straumann, Ueli Stürzinger, Franz Immer

## Abstract

Median organ waiting times published by transplant organizations around the world may be biased when death or censoring is disregarded. This can lead to too optimistic waiting times for all organ waiting lists, but most strikingly in kidney transplantation, and as a consequence, may deceive patients on the waiting list, transplant physicians, and healthcare policy maker. In this cohort study of all transplant candidates in Switzerland, we use competing risk multistate models for the analysis of time-to-event data of the organ waiting list. The resulting cumulative incidences are probabilities for transplantation or death by a given time are a more accurate description of the events occurring on the waiting list. In accordance with the concept of median survival time in survival analysis in clinical trials, we suggest the median time to transplantation (MTT), the waiting time duration at which the transplant probability is 0.50, as a measure of average waiting time.

**Summary points:**

- Transplant candidates on the waiting list, healthcare professionals and policy makers may be interested in the accurate assessment of the probabilities of transplantation and undesirable events.
- The median waiting time based on the median calculated across the waiting durations of transplanted individuals may be misleading due to selection bias, ignoring death and censored observations.
- Competing risk multistate models based on the Aalen-Johansen estimator are effective in assessing the waiting list with multiple competing events.
- From the cumulative incidence curve the probabilities of transplantation or death can be determined for any time period.
- We propose to report both the median time to transplantation (MTT), i.e. the waiting time duration at which the transplant probably is 50%, as well as the probabilities of other competing events during MTT.
- National transplant organizations around the world may consider changing their reporting policy regarding waiting list statistics.

---

Transplant organization sometimes report the average across the waiting times of organ recipients, i.e. the average of the durations from registration on the waiting list until transplantation. Typically, the distribution of waiting times is right-skewed; thus, often the median is used as the central value (see Figure 1). For example, in Switzerland the median waiting time across transplanted persons was 0.52 years for a heart, and 1.13 years for a kidney in a cohort registered and transplanted between 2018 and 2022. Per definition of the median, 50% of the transplanted individuals had shorter, and 50% had longer waiting time than the 0.52 years (heart) or the 1.13 years (kidney).

**Figure 1:**
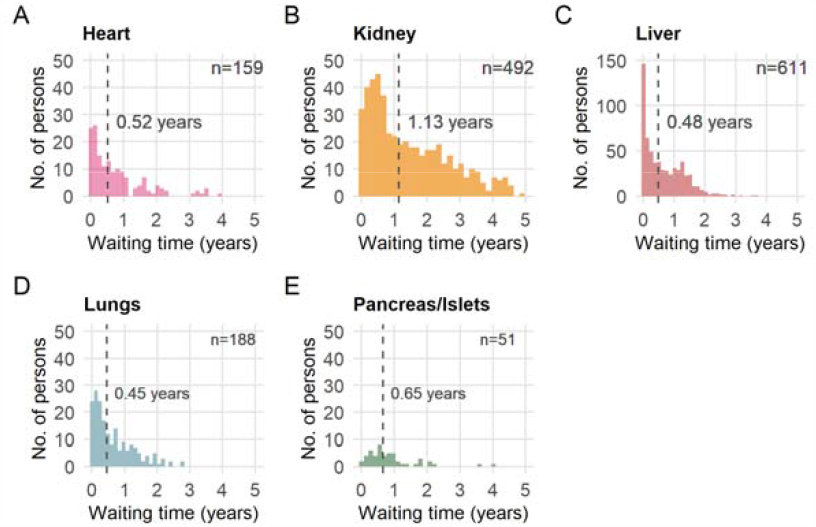
Distribution of waiting times for the different organs A—E (from date of registration to date of transplantation) of 1,501 persons listed after 1. January 2018 and transplanted before 31. December 2022 in Switzerland. The median waiting time is shown as dashed vertical line. Only transplanted persons are considered in such an analysis.

However, there are a number of issues with reporting waiting times in this manner and there are more appropriate methodology from survival and competing risks models available [1,2]. Such alternative strategies for analysing the organ waiting list have long been proposed [3–5], but have not been widely adapted.

The most commonly asked questions by transplant candidates are “How likely is it to receive an organ?” and “How long will I have to wait?”. Unfortunately, the “median waiting time” as described above is not an appropriate summary statistic to answer these questions. There are good arguments that it may be misleading to say that transplant candidates need to wait 0.52 years for a heart, and 1.13 years for a kidney in average.

## Selection bias

Selection bias occurs when patients in a study or analysis differ systematically from the population of interest, leading to a systematic error in the results. The median waiting time is based on transplanted persons only, a sub-population from the population of transplant candidates on the waiting list, and most likely not a very representative one. The allocation of organs is not at random; instead it is on the basis of legally defined criteria, i.e. medical urgency, medical utility, waiting time and equality of opportunity. Thus, the persons on the waiting list and the persons transplanted may be two different populations and the median waiting time cannot simply be generalized to all the individuals on the waiting list.

Even though it may be correct to say, “transplanted persons waited 0.52 years for a heart (or 1.13 years for a kidney) in average”, this is the answer to the wrong question. It is the individuals on the waiting list who want to know how long they have to wait, and not those who already had been transplanted. From the perspective of a person on the waiting list, the statistic is conditioned on the future, i.e. the waiting time is conditional on that a candidate will receive a transplant; however, nobody on the waiting list knows whether they will get a transplant or not. This makes the statistics meaningless from the perspective of a waiting transplant candidate and the medical team in charge.

## Ignoring death and censoring

The median waiting time does not acknowledge that persons on the waiting list may die, or that not everyone will receive and organ transplant. In other words, the statistic would only be valid in a world in which all persons on the waiting list are transplanted, and no one dies (a fictitious world that does not exist). The censored observations are ignored, these are transplant candidates who have not yet experienced an event (transplantation, death or delisting) and are still waiting. Methodology from competing risk multistate models are more appropriate and can effectively address the events occurring on the waiting list by using all the data available.

## Competing risk multistate model

In a multistate model persons can transition from one state to the next. For example, on the transplant waiting list, persons can transition from the waiting list to three states, transplanted, death or delisting (Figure 2). In survival analysis this is known as competing risks, i.e. when other events (competing risks) precludes the occurrence of the primary event (transplantation). For example, death or delisting will exclude a transplantation. Likewise, a transplanted person cannot die anymore on the waiting list. In the example of the waiting list model, only transitions from the initial state to one of the competing risk states are allowed. It would not be possible that a person receives an organ transplant and later would die on the waiting list. Delisting is a third state that may occur, for example, when a person is considered too sick or too old for a transplantation [6].

**Figure 2:**
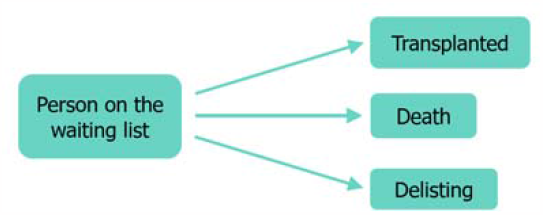
Simple multistate model for the organ waiting list with death and delisting as competing events.

For the estimation of the transition probabilities the Aalen-Johansen estimator can be used; it is an extension of the Kaplan-Meier estimator for a Markov process with a finite number of states. The resulting cumulative incidence curves for each state are very natural to interpret as the probability that an event is occurring in a given period of time. The Aalen-Johansen estimator has the elegant property that the probability of transplantation until time *t*, plus the probability of death until *t*, plus the probability of delisting until *t*, plus the probability of still waiting until *t*, is exactly one.

A recent work [7] proposed a rather inconvenient combination of reporting Kaplan-Meier estimates complementary to cumulative incidence analysis; however, it has been well established that Kaplan-Meier and competing event censoring is not recommended as such an approach provides upward biased estimates of the cumulative incidence [8].

## Data source, cohort and methods

The data source was the Swiss Organ Allocation System (SOAS) which is used by Swisstransplant for national organ allocation and also contains the national waiting list for each organ.

We analysed the complete cohort of transplant candidates in Switzerland listed on 1. January 2018 or later and observed all persons for a 5-years period until 31. December 2022 (end of study date). Persons with planned living donation at the date of registration, or persons who received a living-donor organ (kidney or liver) were excluded from the analysis. Also, observations on the small bowels waiting list were excluded as there were only 3 transplant candidates listed in the observed time period.

The resulting analysis dataset involved N=2,992 transplant candidates with N=3,083 observations due to multi-organ listings (N=91), most commonly kidney-pancreas and liver-kidney. Descriptives of the cohort are shown in Table 1. On the very rate occasion of loss to follow-up, persons were delisted. There was no missing data.

**Table 1:** Descriptives of the transplant candidate cohort.

|  | **Transplant candidates**<br>**on the waiting list**<br>**(N = 2,992)** |
| --- | --- |
| N (persons) | 2,992 |
| n (observations) | 3,083 |
| Date of registration, range | from 01.01.2018<br>to 29.12.2022 |
| Age, median (IQR) | 56.0 (45.0–63.0) |
| Female, n (%) | 1,013 (33.9%) |
| Multi transplant listings, n (%) | 91 (3.0%) |
| Bloodtype n (%) |  |
| A | 1,306 (43.6%) |
| AB | 123 (4.1%) |
| B | 350 (11.7%) |
| O | 1,213 (40.5%) |

We used an incidence cohort, and recipient registerer before 2018 were not considered, even though they may have been transplanted during the observation period. Including these individuals (left truncation) may introduce immortal time bias because they have immortal time until 1. January 2018 and also a type of survivorship bias as persons listed on the same day but had an event (transplantation, death or delisting) before 1. January 2018 would be excluded.

The median follow-up time was 2.3 years (estimated by inverse Kaplan-Meier).

We fitted a multistate model using “survfit” from the “survival” package v3.5-7 [9,10] using the Aalen-Johansen estimator for multi-state survival. For visualizations of the cumulative incidence we used the “ggsurvfit” v1.0.0 package [11]. All analyses were performed using R v4.3.2 [12].

The following transitions were observed: 1,501 transplantations, 245 deaths, and 159 delistings. At the end of study 1,178 transplant candidates were censored.

We reported this study following the guidelines Strengthening the Reporting of Observational Studies in Epidemiology (STROBE) statement [13].

## Probabilities of transplantation, death, delisting, and still waiting

The cumulative incidence functions for the different states are shown in Figure 3. Probabilities of being in each state for 1-year, 2-year, 3-year, and 4-year are shown in Table 2.

**Table 2:** Probabilities (P) of the different states by 1-year, 2-years, 3-years, and 4-years periods on the waiting list.

|  | P(transplanted) | P(still waiting) | P(death) | P(delisting) |
| --- | --- | --- | --- | --- |
| **1-year** |  |  |  |  |
| Heart | 0.47 | 0.40 | 0.09 | 0.04 |
| Kidney | 0.19 | 0.78 | 0.01 | 0.01 |
| Liver | 0.42 | 0.42 | 0.11 | 0.04 |
| Lungs | 0.53 | 0.39 | 0.06 | 0.02 |
| Pancreas/Islets | 0.42 | 0.57 | 0.01 | 0.00 |
| **2-years** |  |  |  |  |
| Heart | 0.60 | 0.22 | 0.12 | 0.06 |
| Kidney | 0.31 | 0.63 | 0.03 | 0.03 |
| Liver | 0.64 | 0.15 | 0.13 | 0.08 |
| Lungs | 0.75 | 0.15 | 0.07 | 0.03 |
| Pancreas/Islets | 0.57 | 0.37 | 0.04 | 0.03 |
| **3-years** |  |  |  |  |
| Heart | 0.64 | 0.16 | 0.13 | 0.07 |
| Kidney | 0.45 | 0.46 | 0.05 | 0.04 |
| Liver | 0.68 | 0.07 | 0.15 | 0.10 |
| Lungs | 0.82 | 0.06 | 0.08 | 0.05 |
| Pancreas/Islets | 0.64 | 0.30 | 0.04 | 0.03 |
| **4-years** |  |  |  |  |
| Heart | 0.76 | 0.03 | 0.13 | 0.08 |
| Kidney | 0.55 | 0.34 | 0.06 | 0.05 |
| Liver | 0.69 | 0.04 | 0.16 | 0.11 |
| Lungs | 0.82 | 0.04 | 0.10 | 0.05 |
| Pancreas/Islets | 0.69 | 0.25 | 0.04 | 0.03 |

**Figure 3:**
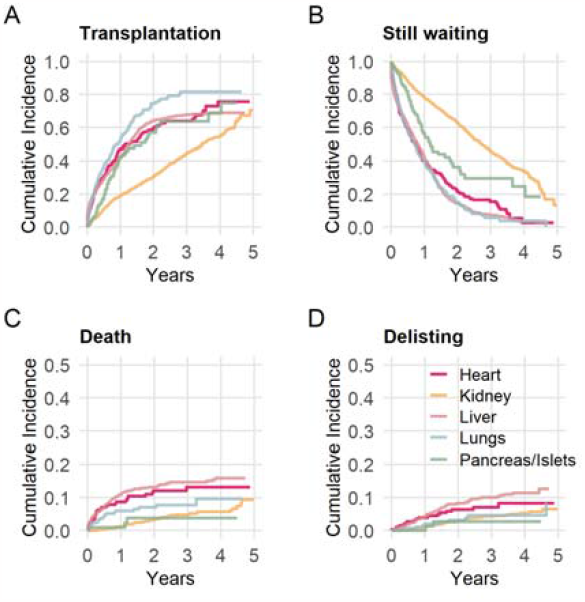
Cumulative incidence curves A—D for the different states of the competing risk multistate model.

For example, the probability of a heart transplant by 2 years is 60%, of death is 12%, of delisting is 6%, and the probability of still waiting is 22% for the heart waiting list. For the kidney waiting list, the probability of transplantation is 31% by 2 years, and 55% by 4 years.

The cumulative incidence of transplantation goes beyond 50% within the first two years after registration on all the organ waiting lists except for kidney which shows a different shape (Figure 3A). After 4 years, the probabilities for waiting for a heart, a liver, or lungs converged close to zero (Figure 3B). Death can occur with a probability of 16% (liver), 13% (heart), 10% (lungs), 6% (kidneys), and 4% (pancreas/islets) during the 4 years after listing (Figure 3C). Delisting is the least common state and occurs most likely on the liver and heart waiting list, with a probability of 11% and 8% by 4 years, respectively (Figure 3D).

The kidney waiting list differs from the other organs. The cumulative incidence of kidney transplantation has a linear shape with the years, and the probability for waiting 4 years (or longer) is still at 34%. There is an apparent increase in the cumulative incidence of death after 4 years on the kidney waiting list, which may need further investigation.

## Median time to transplantation (MTT)

In survival analysis of clinical trials, it is common to report the median survival time to evaluate the efficacy of a novel treatment. It is the length of time corresponding to a probability of 0.50 of surviving this timepoint. Applying this concept to the waiting list, the median time to transplantation (MTT) would correspond to a probability of 0.50 of being transplanted until this timepoint. To illustrate this, we have highlighted the MTT in the cumulative incidence curves (Figure 4). More informally, it is the timepoint by which 50 out of 100 persons on the waiting list are expected to be transplanted; thus, it can indeed be seen as an average waiting time.

**Figure 4:**
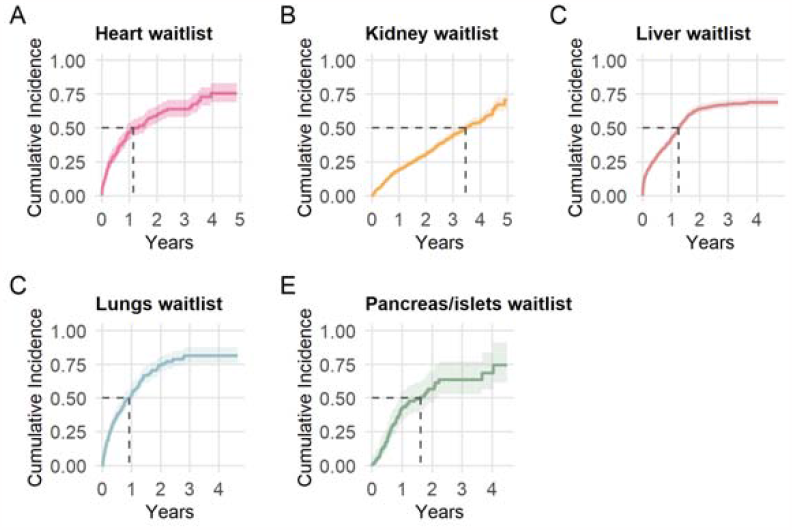
Cumulative incidence curves for transplantation for the different organs A—E with 95% confidence bands and median time to transplantation (MTT) defined as the duration corresponding to the cumulative incidence of 0.50 (dashed line).

A direct comparison of the MTT to the simple median of the waiting times gives quite different results (Table 3). For example, taking the median across all kidney recipients listed in 2018 to 2022 is 1.13 years; however, the MTT is 3.45 years which is a difference by a factor of 3. It may also be important to communicate risks to patients as well, therefore the probability of death during MTT is also provided.

**Table 3:** Direct comparison of the median waiting time versus the median time to transplantation (MTT) based on a competing risk multistate model.

|  | Ignoring competing events and censoring | Competing risk multistate model |  |
| --- | --- | --- | --- |
|  | Median waiting time | Median time to transplantation MTT | Probability of death |
| Population | Transplanted persons only | All persons on the waiting list |  |
| Sample Size | N=1,501 | N=3,083 |  |
| Estimator | Sample median across waiting time durations | Aalen-Johansen |  |
| Interpretation | Half (50%) of transplanted persons had shorter, and 50% had a longer waiting time. | The waiting time duration at which the probability of transplantation is 50%. | The probability of death during MTT. |
| Organ waiting list |  |  |  |
| Heart | 0.52 years | 1.14 years | 9% |
| Kidney | 1.13 years | 3.45 years | 6% |
| Liver | 0.48 years | 1.26 years | 12% |
| Lungs | 0.45 years | 0.92 years | 6% |
| Pancreas/islets | 0.65 years | 1.62 years | 4% |

## Communicating to patients

Communicating to patients on the organ waiting list about their prognosis and potential likelihood of transplantation and adverse outcomes could be challenging. First, it certainly is an emotional subject with large uncertainties. Second, the probabilities are based on groups of people and may be difficult to relate to an individual. Finally, it may be hard to balance between statistical accuracy and simple terms.

When MTT as explained above is not easily understood, and probabilities are unhelpful for patients, it may be more helpful to use frequencies [14]. For example, a clinician could explain to a heart transplant candidate, “If there were 100 people in your situation, 50 are expected to be transplanted by 1.14 years, and 9 may die on the waiting list”. For kidney transplantation, one could say, “for 100 listed kidney transplant candidates, 50 are expected to be transplanted by 3.45 years, but 6 may die on the waiting list”.

## Conclusion

Calculating the sample median of waiting times across transplanted persons is not appropriate as an estimate of the average waiting time to communicate to transplant candidates. The reason is such a simplified approach ignores death and censoring. A number of biases can occur in the process when the analysis is not done carefully: selection bias (only considering transplanted persons), survivorship bias and immortal time bias (when all transplanted individuals are considered as events without a fixed enrolment date).

Results show much longer waiting times when the appropriate analysis is performed which is more in consistence with what clinicians and transplant candidates experience in everyday clinical practice. A limitation of our approach could be that MTT obviously cannot be calculated in countries where the 0.50 probability is not reached in a reasonable amount of years. It is clear that our estimates cannot be generalized to other countries, but we hope to provide convincing reasons so that transplant organizations do not report average wating times without considering death and censored observations appropriately.

Understanding the waiting list as a competing risk multistate model and visualizing the cumulative incidence can inform not only clinicians but also patients when the probability of transplantation and risks are carefully explained, may lead to novel insights about the waiting list and can support the monitoring of national transplant programs.

## Data Availability

The database used are owned by the Federal Office of Public Health (FOPH), Switzerland, which establishes the requirements for their access and use.

## Patient and public involvement

Patients were involved in the conduct of this research. The design of the study, the results and the communication of results were critically discussed with a member of the patient advisory board.

## Contributors

All authors declare to meet the ICMJE condition for authorship. CRediT author statement: SS: Conceptualization, Methodology, Validation, Formal Analysis, Writing – Original Draft, Writing – Review & Editing, Visualization, Project Administration. AE: Conceptualization, Writing – Review & Editing, Supervision. DS: Writing – Review & Editing. LS: Validation, Writing – Review & Editing. US: Writing – Review & Editing. FI: Conceptualization, Writing – Review & Editing, Supervision.

## Ethical approval

The project was submitted to the Ethics Committee of the Canton of Bern which granted an exemption from requiring ethics approval (KEK Bern; 2023-01371). The project does not fall under the Human Research Act; legal regulation in Switzerland follows an internationally applied distinction between research subject to approval and quality assurance that is not subject to approval. The present study meets the criteria for a quality assurance project in terms of reporting of official national statistics of transplant activity and outcomes on the waiting list. According to the Swiss “Organ Allocation Ordinance”, the national allocation centre (Swisstransplant) has the following task (among others): it compiles statistics on the donation, allocation and transplantation of organs (Article 34d lit. c).

